# Institutional Residence Six Months After Acute Ischaemic Stroke: Prognostic Model Development, Internal Validation, and Between-Country Variation in the International Stroke Trial

**DOI:** 10.64898/2026.09.24.26363979

**Authors:** Alex Kubiak, Farzard Haji Boloori, Nicholas Powers, Zhuoyan Wu, Phuc Tran, Olufemi Osikoya

**Author notes:** **Corresponding Author**, Alex Kubiak BSc Department of Surgery, Kansas City University School of Medicine, 1750 Independence Ave, Kansas City, MO 64106, United States.

## Abstract

**BACKGROUND:** Existing models for discharge to long-term institutional care after stroke are developed within single health systems and pool to an area under the curve near 0.80, but none is accompanied by an estimate of how much of the outcome is a property of the health system rather than the patient.

**METHODS:** Secondary analysis of the International Stroke Trial, a randomised trial in 19,435 patients with suspected acute ischaemic stroke recruited in 1991–1996 across 36 countries.

Among 14,885 survivors to six months with residence recorded, we developed and internally validated a logistic prognostic model for residential or nursing-home residence using 17 admission-day terms, and compared it against a pre-specified benchmark of age and stroke syndrome alone. Between-country variation was estimated by a method-of-moments decomposition with a cluster bootstrap.

**RESULTS:** Institutional residence was recorded for 1,984 of 14,885 survivors (13.3%); the model was fitted on 14,121 patients containing 1,935 events. In 10-fold cross-validation with the whole procedure refitted per fold, the out-of-fold c-statistic was 0.789, calibration slope 0.987, calibration-in-the-large 0.1371 predicted against 0.1370 observed, and Brier score 0.1013. A benchmark of age and stroke syndrome alone reached an optimism-corrected 0.774 against 0.790 for the full model, a corrected difference of 0.0160. Between-country variation gave an intraclass correlation of 0.0409 (0.0191 to 0.0673), and in leave-one-country-out validation the c-statistic ranged from 0.635 to 0.823 across the 14 countries with enough events to estimate it.

**CONCLUSIONS:** Institutional residence after stroke can be stratified at admission with discrimination comparable to existing models and calibration that holds in unseen patients, but almost all of the signal is carried by age and stroke syndrome. Performance varies substantially between countries, pointing to placement systems as the limit on what a pooled model can do.

## INTRODUCTION

Discharge to long-term institutional care is among the most consequential outcomes of stroke. It reshapes a patient’s life, it is the outcome families ask about first, and it is a major driver of the long-term cost of stroke to health and social care systems. Yet the proportion of stroke survivors newly admitted to long-term care varies several-fold across published cohorts, from 7% to 39% with a median of 17%, and the model of care provided in the long-term care setting is usually undefined.1

Two explanations compete. The variation may reflect case mix: older and more severely affected populations generate more placements. Or it may reflect the system: the availability of care-home beds, the threshold for admitting to one, and what a country counts as institutional care. A national audit of 79 hospitals attributed regional variation in institutionalisation rates partly to differences in ease of access, asked whether the rate could serve as an indicator of hospital care quality, and concluded that individual-level prediction at admission was not reliable enough to justify early resource-allocation decisions.2

Prediction models for discharge destination after stroke already exist and perform reasonably. A 2025 systematic review of 14 studies and 22 models reported areas under the curve from 0.75 to 0.95 and a pooled estimate of 0.80 (95% CI, 0.75–0.86) across five validation models, while judging every included study to be at high risk of bias.3 An established risk score reached a c-statistic of 0.75 (0.72–0.78) for placement in a care institution among 980 patients in a French registry.4 Three features of that literature limit what it can answer: the models are developed within single health systems, so none can say whether performance transports; the outcome is usually discharge disposition ascertained from hospital coding, which conflates the patient’s state with one system’s coding conventions and bed availability; and none is accompanied by an estimate of how much of the outcome variance sits above the patient.

Distinguishing case mix from system therefore requires a cohort in which eligibility, ascertainment and follow-up are uniform across many countries. The International Stroke Trial provides one.5 Recruitment ran from 1991 to 1996 across 466 deposited hospital codes in 36 countries, with central randomisation, uniform eligibility and follow-up over 99% complete, and the individual patient data are publicly available.6 Because residence at six months was recorded by trial follow-up rather than by hospital discharge coding, the outcome is comparably ascertained across all participating countries. Between-country differences in outcome within this trial have been described before, in an analysis that found case-mix adjustment explained only part of them,7 but the variance has not been partitioned formally for this outcome.

We asked three questions. Can institutional residence at six months be predicted from information available on the day of admission? How much of that prediction requires detailed acute-phase assessment, as opposed to age and stroke syndrome alone? And how much of the variation in the outcome lies between countries rather than between patients?

## METHODS

### Design and source of data

This is a secondary analysis of individual patient data from the International Stroke Trial (IST), a multicentre randomised trial of up to 14 days of aspirin, subcutaneous unfractionated heparin, both, or neither, in patients with suspected acute ischaemic stroke.5 The present analysis is observational; the randomised allocations play no part in the model. This is a prognostic model development study with internal validation, reported according to TRIPOD+AI.8 The completed checklist and a PROBAST self-assessment are offered as supplemental material upon request.

No external validation cohort was analysed.

### Participants and cohort provenance

#### Source population and provenance

Patients with a clinical diagnosis of acute ischaemic stroke, onset within 48 hours, and no clear indication for or contraindication to aspirin or subcutaneous heparin; criteria were inherited in full from the parent trial and not modified.5 Data were collected prospectively on standardised forms and validity-checked at the coordinating centre in Edinburgh, with central randomisation.6 One record is one patient, and the record count equals the published randomised total of 19,435.

#### Cohort for this analysis (Figure 1)

The model is fitted in survivors only. Of 19,435 randomised patients, 4,369 had died before six months and 156 had no recorded six-month vital status; a further 25 survived with residence unrecorded. The two mechanisms are reported separately because they differ in kind: death is structural to the design, whereas unrecorded residence among survivors is potentially informative missingness in the outcome. This left 14,885 survivors with residence recorded; listwise deletion removed a further 764 (5.1%), leaving 14,121 patients with 1,935 events on whom the model was fitted. This is a survivor-conditional model and does not apply to patients who die.

**Figure 1.**
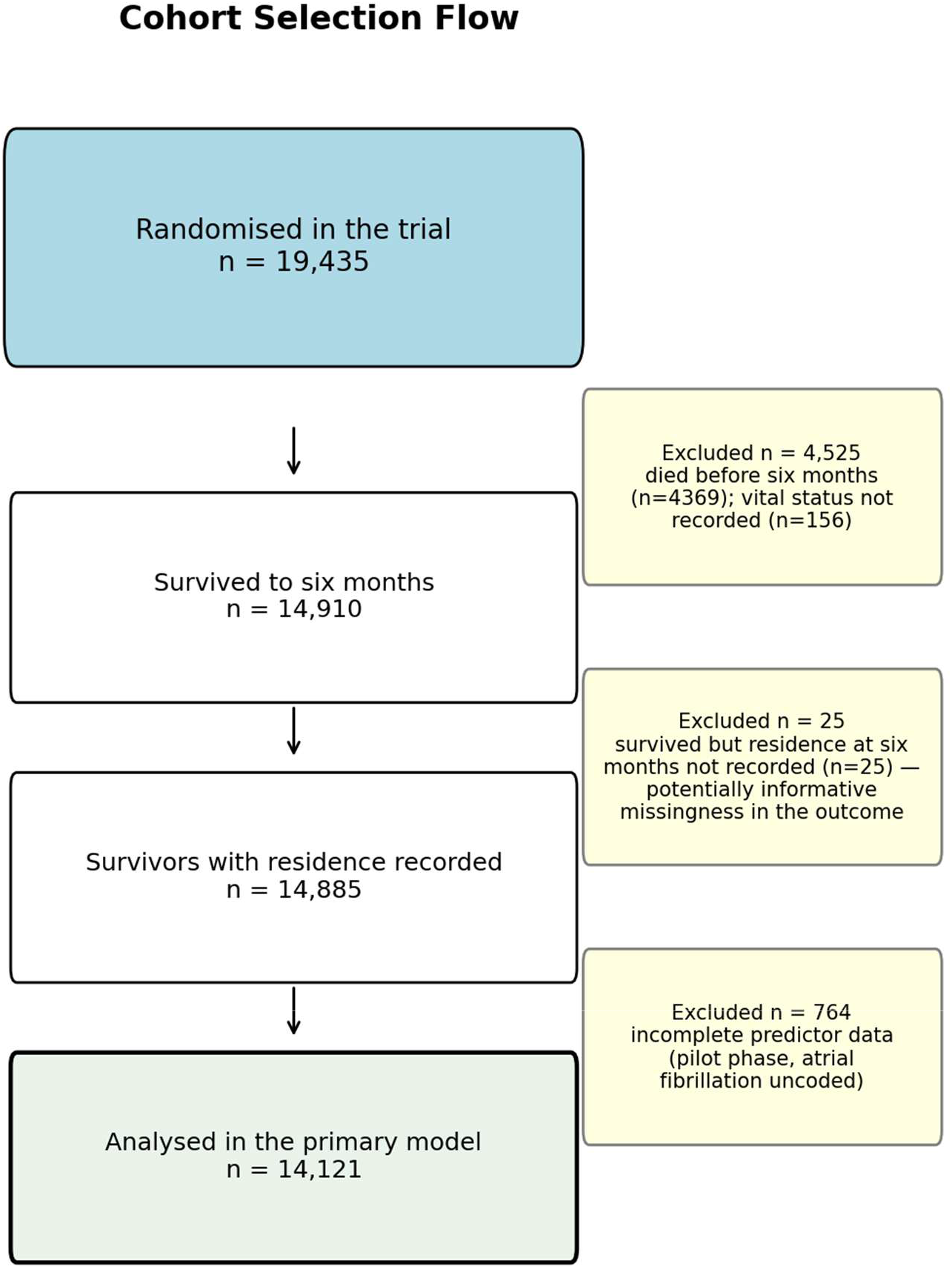
Cohort selection. Three stages are shown, and the two exclusion mechanisms are kept separate because they differ in kind. Of 19,435 randomised patients, 4,369 had died by six months and 156 had no recorded six-month vital status, leaving 14,910 survivors; a further 25 survived but had residence at six months unrecorded, which is potentially informative missingness in the outcome itself, leaving 14,885. The 764 patients removed at the final stage are the pilot-phase patients (1991–1993) for whom atrial fibrillation was never coded, so the primary analysis is effectively restricted to the main trial phase.

#### Outcome

Residential care or nursing home as the recorded place of residence at six-month follow-up, against home or a relative’s home. Ascertainment was by trial follow-up rather than hospital discharge coding, so the outcome is comparably defined across countries at the level of the recorded category. Pre-stroke residence was never recorded, so this is residence at six months, not institutionalisation caused by the stroke.

#### Predictors

Seventeen terms available on the day of randomisation: age as a restricted cubic spline (knots at 55, 68, 78 and 88 years), sex, conscious level, Oxfordshire Community Stroke Project syndrome, systolic blood pressure, atrial fibrillation, a count of eight neurological deficits, and four individual deficit indicators. The set was frozen before any model was fitted.

#### Source-publication reconciliation

Eight integrity checks reproduce the trial publication’s figures exactly, including the randomised total, 14-day deaths in each treatment arm, the six-month dead-or-dependent total and all three final-diagnosis strata. Two quantities do not reconcile and are reported rather than resolved: 466 deposited hospital codes against 467 reported, and no cut-point on the deposited age variable that reproduces the published proportion aged over 80.

### Data source and ethics

International Stroke Trial database, version 2 (corrected public-use file), Edinburgh DataShare, https://doi.org/10.7488/ds/104, under the Open Data Commons Attribution Licence. The data are anonymised and publicly deposited; no institutional review board approval or informed consent was required.

### Statistical analysis

A logistic regression model was fitted to the 17 pre-specified terms. Discrimination is the c-statistic, corrected for optimism by 500 bootstrap replicates in which the full modelling procedure was repeated in each resample, the method with the least bias for internal validation of a logistic model.9 Calibration is reported as the optimism-corrected calibration slope. Success was pre-specified as an optimism-corrected c-statistic of at least 0.75 with a calibration slope between 0.85 and 1.15.

Which performance metrics are informative. Two reported quantities are constrained by the fitting procedure rather than estimated from it: the apparent calibration slope is 1.000 by construction, and apparent calibration-in-the-large equals the observed event fraction by construction, because a logistic model with an intercept forces mean predicted probability to equal observed prevalence in its own sample. The apparent Brier score and grouped calibration plot are similarly optimistic. None is evidence that the model is calibrated, and we label them apparent rather than omitting them. Calibration-in-the-large, the calibration slope, the Brier score, the calibration curve and the decision curve are therefore additionally computed out of fold, from 10-fold cross-validation in which every coefficient is refitted inside each fold and predictions generated only for held-out patients.

### Clinical usefulness

A decision curve plots net benefit against threshold probability.10 The clinical action assumed at threshold is early referral to social work and care-placement planning during the acute admission — a low-cost, reversible action, which is why a threshold range of 0.05 to 0.30 was chosen rather than a default 0 to 0.50: below 5% the action would be taken for almost everyone, and above 30% the model rarely assigns risk in this cohort.

### Benchmark comparison

A model containing age and stroke syndrome alone was pre-specified as a comparator, to test whether the full model represents more than repackaged age and severity, and is fitted on the same complete-case rows. Confidence intervals for each c-statistic and for their difference come from a paired bootstrap of 500 replicates with both models refitted within each replicate and evaluated on the same resampled patients. Because a 17-term model carries more optimism than a 2-term one, the difference is additionally optimism-corrected by a Harrell bootstrap applied to both arms. The benchmark is a discrimination comparison only; it was not separately calibrated and is **not** offered as a validated clinical rule.

### Between-country variation

Variance between countries was decomposed by a method of moments, subtracting the variance expected from binomial sampling at the observed cluster sizes from the observed variance of country-specific rates, expressed as an intraclass correlation on the probability scale.11,12 One country contributing fewer than five patients was excluded, leaving 35; intervals come from resampling countries, not patients, with 1000 replicates. A random-intercept mixed model returned a variance component of exactly zero with a singular covariance which is a boundary artifact previously demonstrated in this dataset and is reported for transparency, not interpreted.

### Internal-external validation

The model was additionally refitted on all other countries and evaluated on each held-out one in turn, giving an out-of-sample c-statistic and calibration slope per country; a country was estimable if it contributed at least 15 events.

### Deviations from the analysis plan

Two were recorded before the analysis was run. First, the plan specified a predictor set including variables recorded at 14 days; the admission-day set was used instead because the discharge variables are missing for 41.3% of survivors, that missingness is associated with the outcome (p < 0.001), and the patients it removes are disproportionately those not discharged alive by day 14. The discharge-augmented model is reported as a sensitivity analysis only. Second, the Fine–Gray model specified for the competing-risk analysis raises NotImplementedError in the analysis stack, so a proportional-odds model on an ordered three-level outcome was substituted. That changes the estimand as well as the estimator: it describes the ordering of outcomes across the whole cohort rather than a subdistribution hazard, so it addresses whether survivor conditioning generates the association but gives no competing-risks estimate of absolute placement risk.

### Missing data

Complete-case analysis was used; 764 patients (5.1%) were dropped from the 14,885 which are the pilot-phase patients recruited in 1991–1993 for whom atrial fibrillation was never coded, a documented feature of the trial rather than data loss, so every primary analysis is effectively restricted to the main trial phase.6 They were younger than those analysed (68.3 versus 70.2 years) and their missingness is associated with the outcome (p < 0.001), so the mechanism should not be assumed ignorable. Multiple imputation (m = 20) was pre-specified; applying the pooled coefficients to the observed design matrix gives a c-statistic of 0.791 and a calibration slope of 1.005, matching complete case.

No formal sample-size calculation was performed, as is appropriate for a fixed cohort; events per variable were 113.8 (1,935 events, 17 terms), exceeding the minimum required by the criteria of Riley et al.13 Analyses used Python 3.12.11 with statsmodels under a single global seed, 20260730.

## RESULTS

### Participants

Of 19,435 randomised patients, 14,885 survived to six months with residence recorded; 1,984 (13.3%) were in residential care or a nursing home. The model was fitted on 14,121 patients with complete predictor data, containing 1,935 events (Figure 1). Those in institutional residence were substantially older (78.10 versus 68.87 years, standardised mean difference 0.825), more often female (61.8% versus 42.2%, 0.400), more often drowsy or unconscious, and had more recorded deficits (3.56 versus 3.06, 0.404) (Table 1).

**Table 1.**
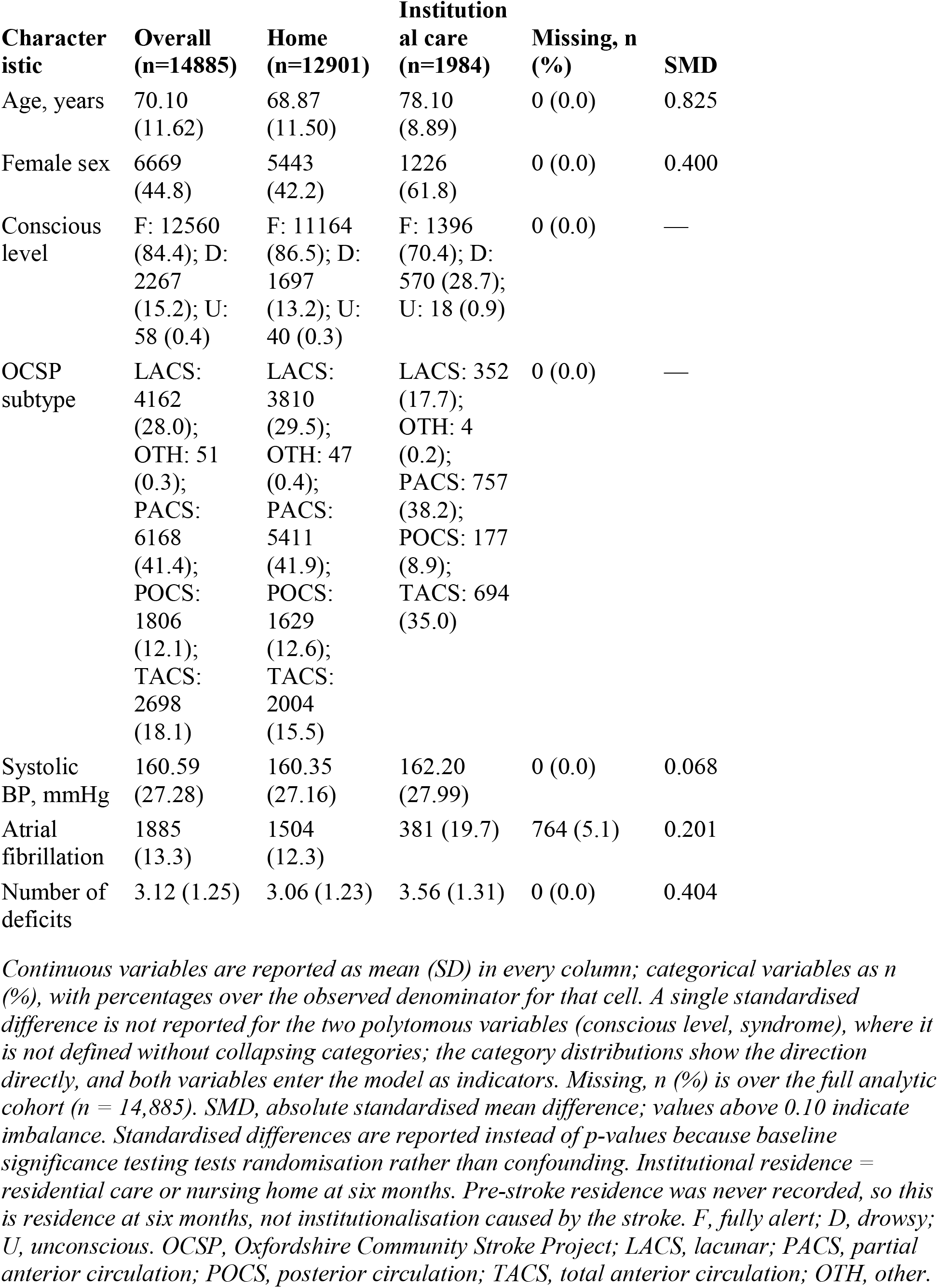
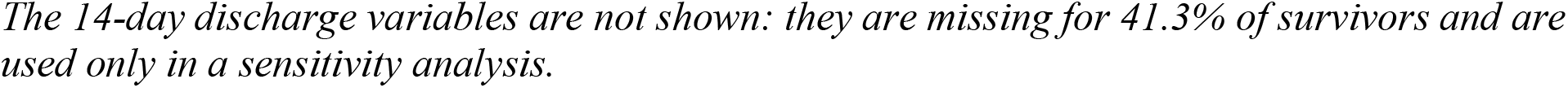
Baseline characteristics by residence at six months.

### Model and performance

Model coefficients are in Table 2. The largest categorical odds ratios were for total anterior circulation syndrome (2.10; 95% CI, 1.69–2.62), drowsiness (1.84; 1.61–2.10), female sex (1.42; 1.27–1.58) and leg or foot deficit (1.42; 1.18–1.73). Unconsciousness carried the largest point estimate (2.87; 1.55–5.31) but rests on 58 patients and 18 events, so it should not be called the strongest predictor. Age contributed non-linearly across its three spline terms; systolic blood pressure did not contribute independently. Atrial fibrillation was commoner among those in institutional residence unadjusted (19.7% versus 12.3%) but its adjusted odds ratio was 0.90 (0.78–1.03), a reversal consistent with confounding by age.

**Table 2.** Prognostic model coefficients.

| Term | OR (95% CI) | p |
| --- | --- | --- |
| AGE_RCS1 | 1.03 (1.01–1.05) | 0.004 |
| AGE_RCS2 | 1.16 (1.10–1.21) | <0.001 |
| AGE_RCS3 | 0.64 (0.54–0.77) | <0.001 |
| RSBP | 1.00 (1.00–1.00) | 0.244 |
| RATRIAL_Y | 0.90 (0.78–1.03) | 0.123 |
| DEFICIT_COUNT | 1.13 (1.03–1.23) | 0.011 |
| RDEF2_Y | 0.98 (0.77–1.24) | 0.854 |
| RDEF3_Y | 1.42 (1.18–1.73) | <0.001 |
| RDEF4_Y | 0.85 (0.73–0.99) | 0.034 |
| RDEF6_Y | 1.14 (0.95–1.37) | 0.158 |
| RCONSC_D | 1.84 (1.61–2.10) | <0.001 |
| RCONSC_U | 2.87 (1.55–5.31) | <0.001 |
| STYPE_OTH | 1.56 (0.52–4.70) | 0.432 |
| STYPE_PACS | 1.26 (1.06–1.50) | 0.009 |
| STYPE_POCS | 1.19 (0.93–1.52) | 0.165 |
| STYPE_TACS | 2.10 (1.69–2.62) | <0.001 |
| SEX_F | 1.42 (1.27–1.58) | <0.001 |

*Fitted on n = 14,121 with 1,935 events. Odds ratios are PREDICTIVE associations; no* *coefficient is interpreted causally, and p-values are shown for completeness rather than for* *inference. Age enters as a restricted cubic spline with knots pre-specified at 55, 68, 78 and 88* *years, so its three terms have no individual clinical interpretation and should be read together.* *RCONSC\_D, drowsy; RCONSC\_U, unconscious (n = 58, 18 events); STYPE, Oxfordshire* *Community Stroke Project syndrome; RDEF2, arm or hand deficit; RDEF3, leg or foot deficit;* *RDEF4, dysphasia; RDEF6, visuospatial disorder; RSBP, systolic blood pressure; RATRIAL\_Y,* *atrial fibrillation present; SEX\_F, female.*

*Fitted on n = 14,121 with 1,935 events. Odds ratios are PREDICTIVE associations; no* Performance is in Table 3. The apparent c-statistic was 0.791 and the optimism-corrected c-statistic 0.790 (n = 14,121), above the pre-specified threshold of 0.75; optimism was 0.002, which is what a 17-term model fitted to 1,935 events should show. The optimism-corrected calibration slope was 0.990, inside the pre-specified window of 0.85 to 1.15. Nagelkerke R^2^ was 0.228.

**Table 3.**
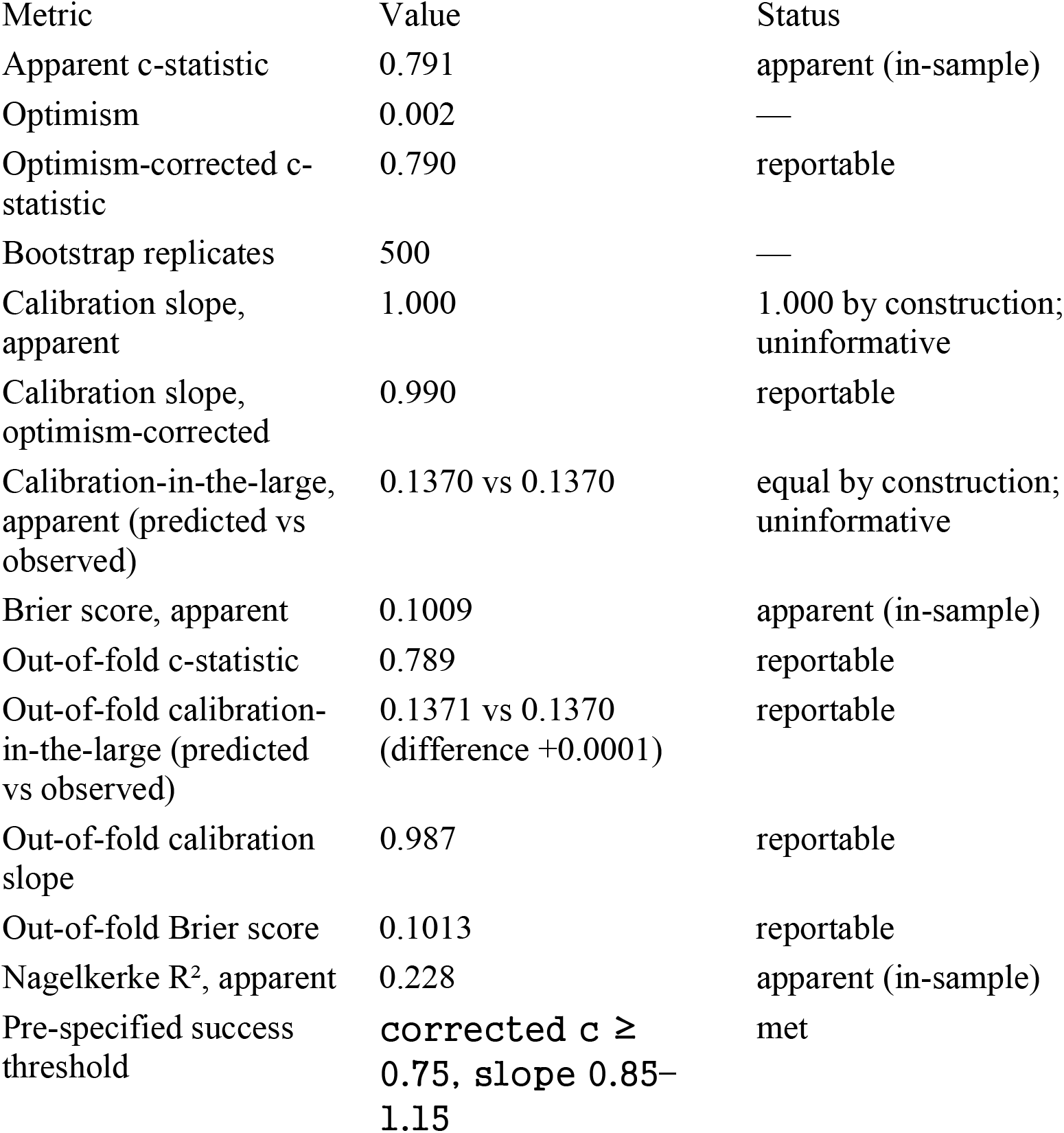
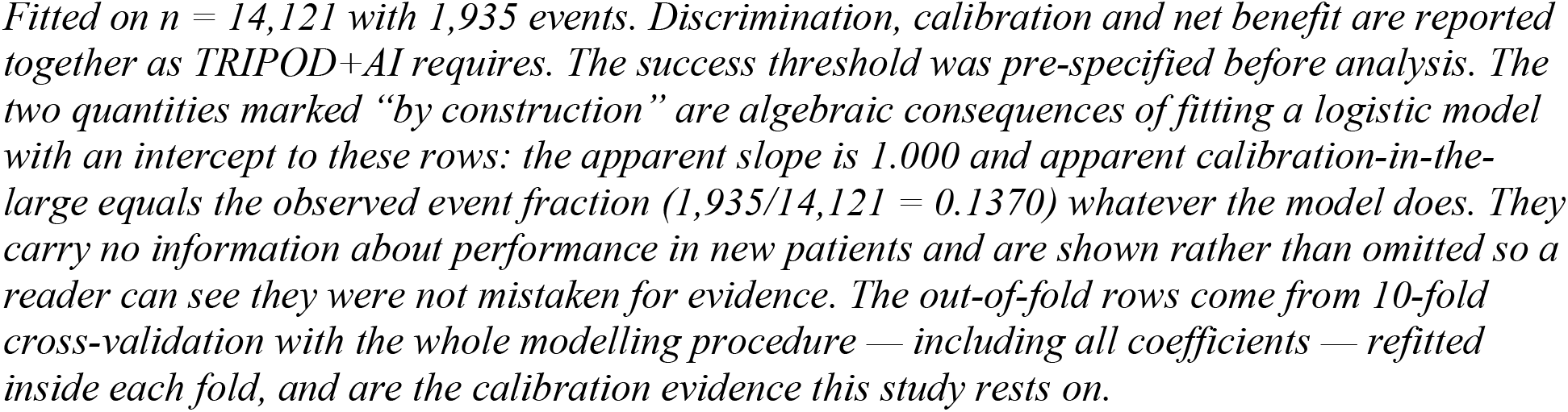
Model performance.

The apparent calibration slope of 1.000 and calibration-in-the-large of 0.1370 against 0.1370 are fixed by the estimation procedure and carry no information. The informative quantities come from 10-fold cross-validation with the whole procedure refitted inside each fold. Out of fold, the c-statistic was 0.789, the calibration slope 0.987, calibration-in-the-large 0.1371 predicted against 0.1370 observed, and the Brier score 0.1013 — essentially unchanged from the apparent values, which is what a 17-term model on 1,935 events should give but is not guaranteed in advance. Figure 2 shows both: apparent deciles, which cannot fall far from the diagonal, and a smoothed curve on continuous out-of-fold predictions, which can.

**Figure 2.**
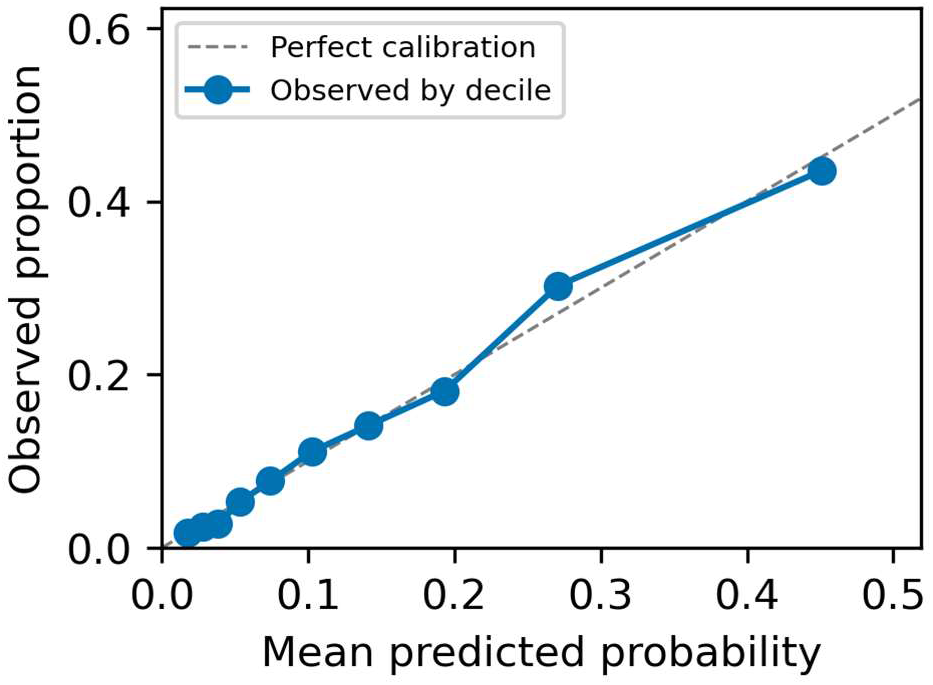
Calibration of the institutional-residence prognostic model. n = 14,121 survivors analysed. Two curves are shown. The decile points are apparent (in-sample) and cannot fall far from the diagonal, because the model was fitted on these rows. The smoothed line is a local-linear fit to continuous out-of-fold predictions from 10-fold cross-validation and is the calibration evidence this study rests on. Optimism-corrected c-statistic 0.790; out-of-fold c-statistic 0.789.

### Clinical usefulness

The decision curve, computed from out-of-fold predictions, is shown in Figure 3. Across threshold probabilities from 0.05 to 0.30 the model gave positive net benefit and exceeded both treat-all and treat-none throughout: 0.0744 at a threshold of 0.10, 0.0549 at 0.15 and 0.0418 at 0.20, against 0.0411, −0.0153 and −0.0787 for treating everyone. Treat-all becomes worse than treat-none above about 0.14, the outcome prevalence. For the assumed action — early referral to social work and placement planning during the acute admission — using the model to select patients is therefore preferable to referring everyone or no one anywhere in the plausible range.

**Figure 3.**
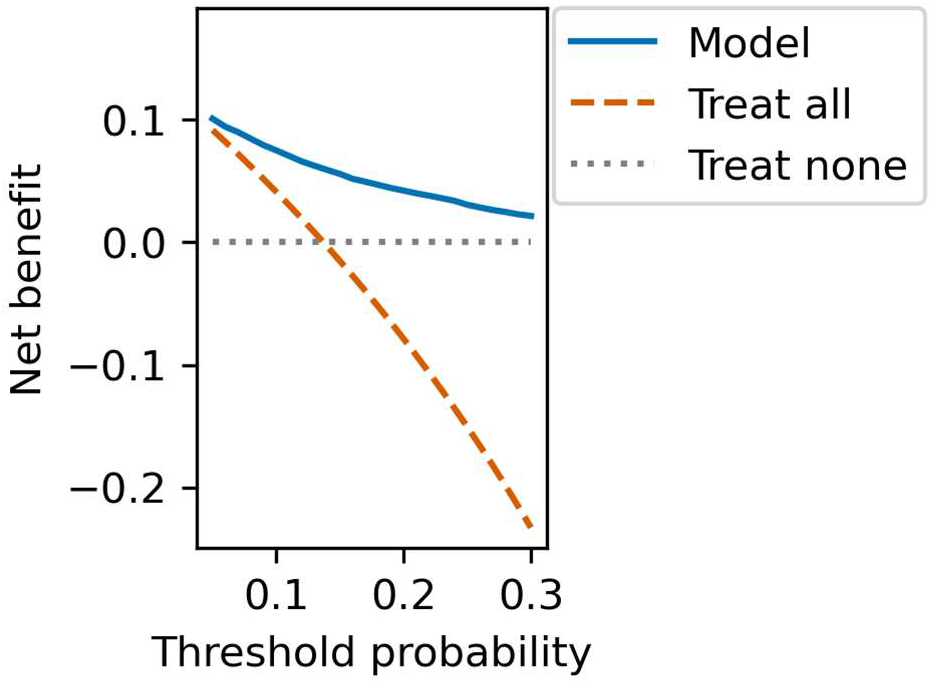
Decision curve for the institutional-residence model, computed from out-of-fold predictions. n = 14,121 survivors analysed. Net benefit is a point estimate and no intervals are shown. Prevalence of institutional residence in the fitted cohort, 0.1370. The assumed clinical action at threshold is early referral to social work and care-placement planning during the acute admission.

### How much of the model is age and syndrome

The pre-specified benchmark containing age and Oxfordshire Community Stroke Project syndrome alone was fitted on the identical 14,121 complete-case rows, so the comparison is a model contrast rather than a mixture of model and cohort. Its apparent c-statistic was 0.774 (95% CI, 0.763 to 0.784) against 0.791 (0.781 to 0.802) for the 17-term model, an apparent difference of 0.0173 (0.0131 to 0.0212) which excludes zero (Table 4); intervals come from a paired bootstrap of 500 replicates with both models refitted and evaluated on the same resampled patients. Optimism-correcting both arms gives 0.790 against 0.774, a corrected difference of 0.0160, the larger model does carry more optimism (0.0017 against 0.0004), but only 0.0013 of the apparent gap was attributable to it. Fifteen additional terms therefore buy a real but small increment. Restricting the full model to patients discharged home gave c = 0.710 (n = 6,331).

**Table 4.**
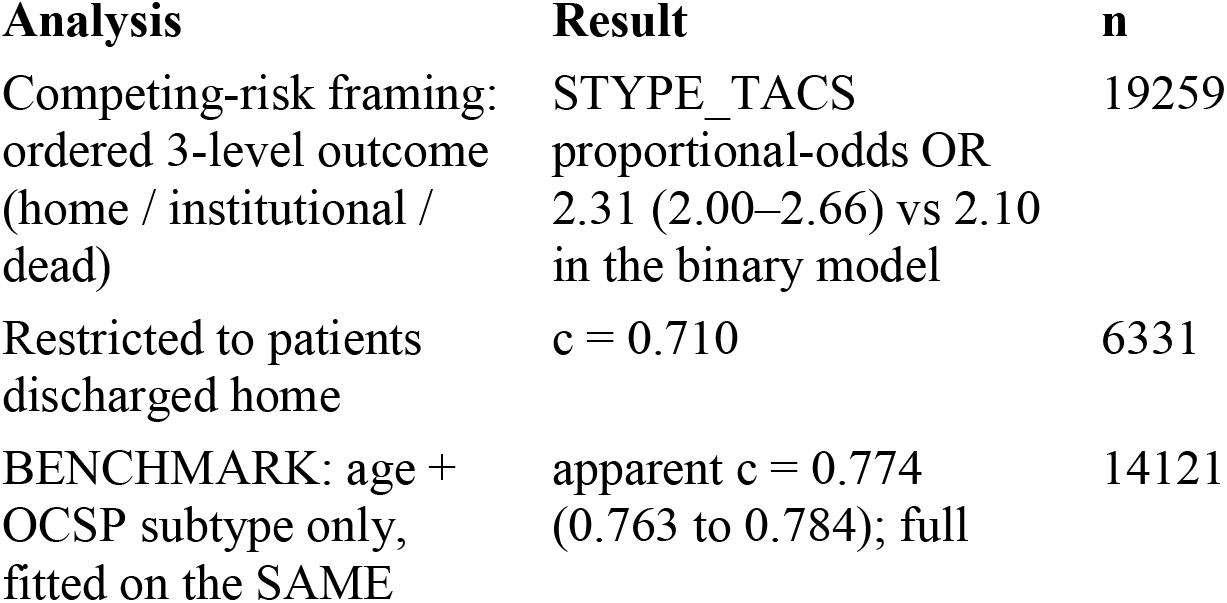
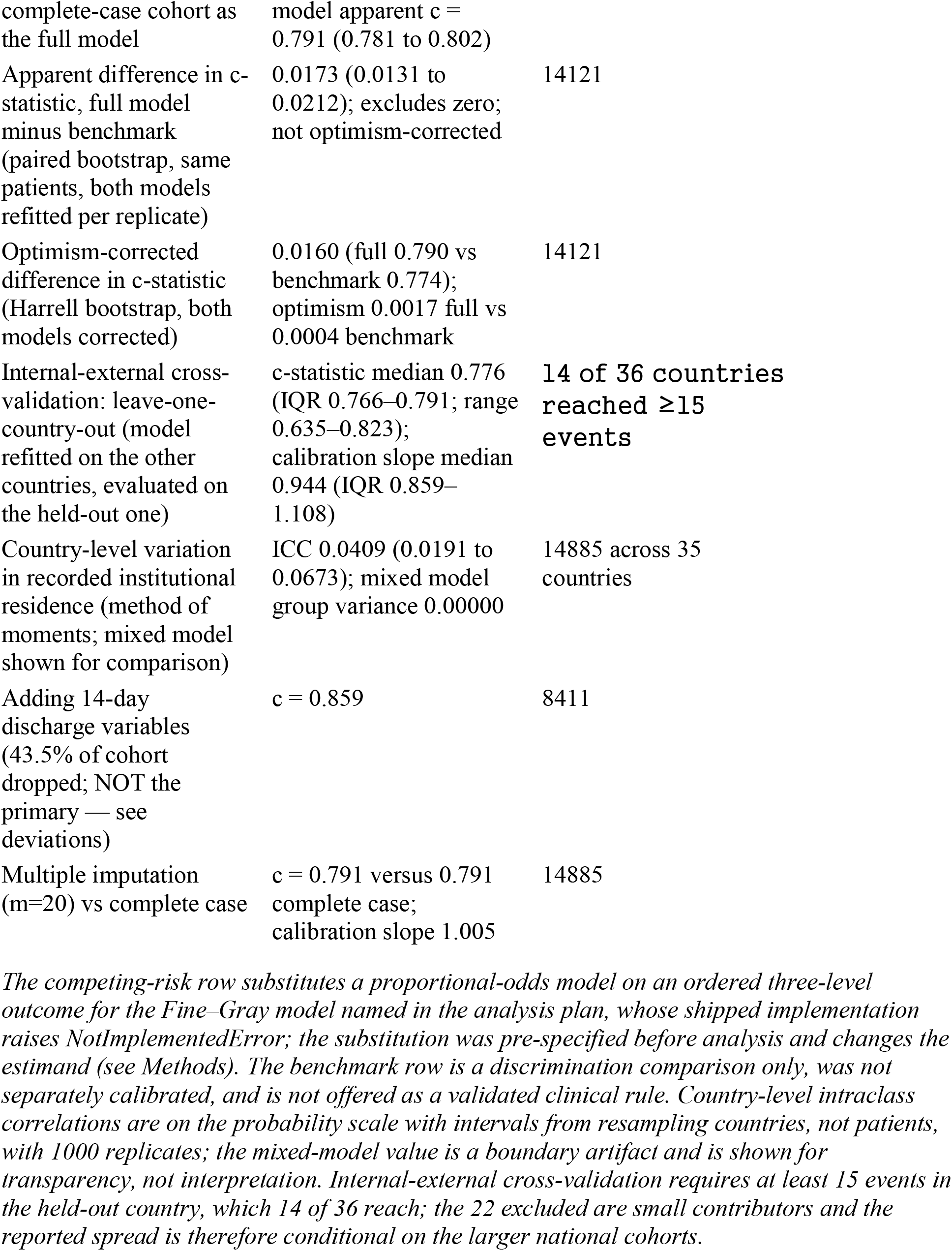
Pre-specified sensitivity and secondary analyses.

Adding the 14-day discharge variables raised discrimination to 0.859, but on 8,411 patients disproportionately excluding those not discharged alive by day 14, whose institutional-residence rate is about double so it is not comparable with the primary and is not offered as a better model.

The two variables carrying that signal show the gradient directly: observed institutional residence ranged from 8.5% after a lacunar syndrome to 25.7% after a total anterior circulation syndrome, and from 3.4% below 65 years to 37.4% at 85 years or above.

### Between-country variation

The method-of-moments decomposition gave a between-country intraclass correlation of 0.0409 (95% CI, 0.0191–0.0673) across 35 countries, excluding zero (Table 4): approximately 4% of the variance in recorded institutional residence at six months lies between countries rather than between patients. The random-intercept mixed model returned a variance component of exactly 0.00000 with a singular random-effects covariance, a boundary artifact of the optimiser rather than evidence of absent variation, reported for transparency only.

Treating death as the worst category of an ordered three-level outcome across all 19,259 patients with a recorded outcome preserved the severity gradient (proportional-odds ratio for total anterior circulation syndrome 2.31, 2.00–2.66, against 2.10 in the binary model), so conditioning on survival does not generate the association.

## DISCUSSION

Institutional residence six months after acute ischaemic stroke was predictable from information available on the day of admission, with an optimism-corrected c-statistic of 0.790 and an optimism-corrected calibration slope of 0.990. Two findings qualify that headline. Almost all of the discriminatory signal was carried by two variables, age and stroke syndrome, which reached an apparent 0.774 between them on the same patients. And a measurable share of the variance in the outcome — an intraclass correlation of 0.0409, excluding zero — lay between countries rather than between patients.

### Prediction at admission

A national audit of 2,778 stroke admissions across 79 hospitals concluded that institutionalisation could not be predicted reliably enough at admission to justify early resource-allocation decisions.2 Our result qualifies that: out-of-fold discrimination of 0.789 with positive net benefit across the plausible threshold range is enough to stratify a population. It is not a correction of the underlying clinical caution — a c-statistic of 0.79 supports service planning and conversations about likely trajectory, not an individual placement decision on day one, and the earlier authors were right about that.

Our discrimination is close to the pooled 0.80 reported across validation models for discharge disposition,3 and above the 0.75 achieved by an established risk score applied to institutional care in a single registry.4 We do not claim a better model, and the comparison is imprecise: those models predict discharge disposition rather than residence at six months, most use severity scales unavailable here, and the pooled estimate comes from studies judged uniformly at high risk of bias.3 What this cohort adds is not performance but the conditions of measurement — uniform eligibility, uniform follow-up ascertainment, 36 health systems — which is what makes the second and third findings possible.

### Two variables carry the signal

The pre-specified benchmark is the more useful finding for practice. Age and stroke syndrome alone gave an apparent 0.774 (0.763 to 0.784); fifteen further admission-day variables added an apparent 0.0173 (0.0131 to 0.0212), an increment that is real but smaller once optimism is accounted for. This is consistent with the systematic-review evidence, which found age and stroke severity to be the two consistently positive predictors across 18 studies and 32,139 participants, and noted that potentially modifiable factors were rarely examined.1

Two readings follow, pointing in opposite directions. Optimistically, a bedside estimate needing only age and syndrome performs almost as well as a 17-term model requiring a calculator, which matters for whether any of this is usable outside a research dataset. Cautiously, such a model is close to a restatement of what a clinician already knows, and its value over unaided judgement was untested here because no clinician predictions were recorded. The increment is small but not zero, so the full model is not merely a repackaging.

### The country-level finding

An intraclass correlation of 0.0409 is not large, but it excludes zero and is larger than the between-country variance in six-month mortality estimated in the same cohort by the same method (ICC 0.0251; 95% CI, 0.0141–0.0370; companion analysis, under review). That ordering is coherent: death is a hard endpoint recorded the same way everywhere, whereas “residential care or nursing home” is a health-system category whose availability, admission threshold and definition differ across countries. An earlier analysis of between-country outcome differences in this trial reached a compatible conclusion, finding case-mix adjustment removed only part of the variation,7 and the national audit reached it within one country.2

Leave-one-country-out validation makes that heterogeneity concrete. Refitting on the other countries and evaluating on the held-out one, the c-statistic had a median of 0.776 (IQR 0.766– 0.791) but ranged from 0.635 to 0.823 across the 14 countries contributing at least 15 events, with calibration slope median 0.944 (IQR 0.859–1.108). The pooled 0.789 is an average over that spread, and in the weakest health system the model is close to useless for a clinical decision. The two country counts here are not interchangeable: the variance decomposition uses 35, having dropped one with fewer than five patients, while cross-validation was attempted in all 36 and returned an estimate for 14.

This has three consequences. It bounds transportability quantitatively rather than in principle: a country adopting this model should expect performance somewhere in that range, not at the pooled value. It partly explains why patient-level predictors plateau, since variance at the country level is unreachable by any patient-level predictor set. And it is a finding in its own right, of the kind the hospital-profiling literature has reached about its own measures: variation in an administratively defined outcome should not be read as variation in the underlying patient state.14,15 For policy, institutionalisation rate is therefore a poor candidate for a cross-national quality indicator without explicit adjustment for placement capacity and for what each system counts as institutional care — the question the original audit asked and left open.2

## Limitations

### The model is survivor-conditional and this is its most important limitation

It was fitted in patients who survived to six months with residence recorded, so it does not apply to patients who die, and 4,550 randomised patients are outside its scope: a clinician applying it must already believe the patient will survive. The competing-risk analysis shows the severity gradient survives when death is included as the worst outcome category, which addresses whether conditioning generates the association but not who the model serves.

### Pre-stroke residence was never recorded

The outcome is residence at six months, not new institutionalisation; some patients were in residential care before the stroke and the model cannot distinguish them. This is the principal reason our 13.3% is not directly comparable with the 7% to 39% range reported for new admissions to long-term care.1

### Calibration is established within this cohort, not beyond it

The out-of-fold slope of 0.987 and calibration-in-the-large of 0.1371 against 0.1370 come from patients the model did not see, so they are evidence rather than arithmetic. But the folds are drawn at random from a pooled 36-country cohort, and the leave-one-country-out slopes range more widely (IQR 0.859–1.108). Calibration in a specific health system should be assumed to need recalibration.

### Internal-external, not external, validation, and no graded severity measure

Leave-one-country-out cross-validation shows how performance travels between the health systems in this trial, but all contributed to development and all are pre-1996; only 14 of 36 countries reached the 15 events needed for a stable held-out estimate, so the reported spread describes the larger national cohorts. Severity is eight binary deficits plus a three-level conscious state rather than a validated scale, which is known to limit stroke outcome models — adding one to a comorbidity-only model for stroke mortality raises discrimination from 0.75 to 0.8216 — so the performance reported here is a floor.

### The cohort predates modern stroke care

IST recruited before intravenous thrombolysis, endovascular therapy or widely established stroke-unit care.17 Organised care changes both survival and function,18 so the mapping from admission-day severity to six-month residence in a contemporary system may differ. The country-level result is likely the more durable finding, since placement systems have not converged.

### Missing data are structural in origin but not demonstrably ignorable

The 5.1% dropped are pilot-phase patients with atrial fibrillation uncoded by design; they were younger than those retained and their missingness is associated with the outcome, though multiple imputation reproduced both the coefficients and the c-statistic. Separately, the discharge-augmented model reaching 0.859 does so on a cohort from which the sickest 43.5% have been removed and should not be quoted as this study’s performance.

Finally, this analysis was planned without formal clinical face-validity review; design questions were addressed by literature synthesis rather than by a practising stroke physician.

## CONCLUSIONS

Among survivors of acute ischaemic stroke, residence in institutional care at six months was predictable from admission-day information with an out-of-fold c-statistic of 0.789 and out-of-fold calibration close to exact, comparable to existing models developed within single health systems. Age and stroke syndrome alone reached 0.774 on the same patients, the other fifteen variables adding 0.0160 after optimism correction. Roughly 4% of the outcome variance lay between countries rather than between patients, and held-out performance ranged from 0.635 to 0.823 across national cohorts. Prediction of institutional residence after stroke is therefore both easier and less portable than it appears: easier, because two bedside variables carry nearly all the signal; less portable, because how well the model works depends on which placement system it is used in.

## Data Availability

The data used in this study were obtained from the International Stroke Trial (IST), a previously completed multicenter randomized controlled trial. The data are not publicly available and are subject to the data-access and governance policies of the original trial. Requests for access to the underlying IST data should be directed to the appropriate data custodian of the International Stroke Trial.

https://doi.org/10.7488/ds/104

## DATA AVAILABILITY

The individual patient data analysed are publicly available from Edinburgh DataShare at https://doi.org/10.7488/ds/104 under the Open Data Commons Attribution Licence: Sandercock P, Niewada M, Członkowska A. International Stroke Trial database (version 2) [dataset].

University of Edinburgh, Department of Clinical Neurosciences; 2011. The full analysis script, the helper library it calls, and the file that reproduces the analytic dataset from the public deposit are archived in a public repository with a citable DOI and are offered as supplemental material.

## RELATED WORK BY THESE AUTHORS

This analysis is one of a series of independent secondary analyses of the International Stroke Trial deposit addressing distinct questions. The between-country intraclass correlation for six-month mortality quoted in the Discussion (0.0251; 95% CI, 0.0141–0.0370) comes from a companion analysis of between-hospital variation in mortality, currently under review.

## ACKNOWLEDGMENTS

Data source: International Stroke Trial database (version 2), Edinburgh DataShare, https://doi.org/10.7488/ds/104, made available under the Open Data Commons Attribution Licence. The IST was funded principally by the UK Medical Research Council, the UK Stroke Association and the European Union BIOMED-1 programme. We acknowledge the International Stroke Trial Collaborative Group, the thousands of patients who joined the trial and their doctors, the Neurosciences Trials Unit which coordinated the study, and the Clinical Trial Service Unit in Oxford which provided randomisation. The depositors bear no responsibility for this secondary analysis.

